# Baloxavir marboxil use for critical human infection of avian influenza A H5N6 virus

**DOI:** 10.1101/2023.09.03.23294799

**Authors:** Wenda Guan, Rong Qu, Lihan Shen, Kailin Mai, Weiqi Pan, Zhengshi Lin, Liping Chen, Ji Dong, Jiawei Zhang, Pei Feng, Yunceng Weng, Minfei Yu, Peikun Guan, Jinchao Zhou, Chuanmeizi Tu, Xiao Wu, Yang Wang, Chunguang Yang, Yun Ling, Sheng Le, Yangqing Zhan, Yimin Li, Xiaoqing Liu, Heyan Zou, Ziqi Huang, Hongxia Zhou, Qiubao Wu, Wenjie Zhang, Jiayang He, Teng Xu, Nanshan Zhong, Zifeng Yang

**Author notes:** W.G., R.Q., L.S., K.M., W.P. and Z.L. contributed equally to this work. Corresponding authors: N.Z. and Z.Y. contributed equally to this work. Zifeng Yang National Clinical Research Center for Respiratory Disease, State Key Laboratory of Respiratory Disease, Guangzhou Institute of Respiratory Health, the First affiliated Hospital of Guangzhou Medical University, 151 Yanjiang Road, Guangzhou, Guangdong, 510120, China., Nanshan Zhong, National Clinical Research Center for Respiratory Disease, State Key Laboratory of Respiratory Disease, Guangzhou Institute of Respiratory Health, the First affiliated Hospital of Guangzhou Medical University, 151 Yanjiang Road, Guangzhou, Guangdong, 510120, China. Author for backup contact before publication, Weiqi Pan, National Clinical Research Center for Respiratory Disease, State Key Laboratory of Respiratory Disease, Guangzhou Institute of Respiratory Health, the First affiliated Hospital of Guangzhou Medical University, 151 Yanjiang Road, Guangzhou, Guangdong, 510120, China. Author contributions Conceived study: Zifeng Yang, Nanshan Zhong Designed study and experiments: Wenda Guan, Rong Qu, Lihan Shen, Kailin Mai, Weiqi Pan, Zhengshi Lin, Zifeng Yang, Nanshan Zhong Performed experiments: Wenda Guan, Rong Qu, Lihan Shen, Kailin Mai, Weiqi Pan, Liping Chen, Ji Dong, Jiawei Zhang, Pei Feng, Yunceng Weng, Minfei Yu, Peikun Guan, Jinchao Zhou, Chuanmeizi Tu, Xiao Wu, Chunguang Yang, Yun Ling, Sheng Le, Yangqing Zhan, Yimin Li, Xiaoqing Liu, Heyan Zou, Ziqi Huang, Hongxia Zhou, Qiubao Wu, Wenjie Zhang, Jiayang He, Teng Xu Interpreted data: Kailin Mai, Weiqi Pan, Zhengshi Lin, Wenda Guan, Rong Qu, Lihan Shen, Nanshan Zhong, Zifeng Yang.

## Abstract

**Background:** Recent increase in human infections of highly pathogenic avian influenza H5N6 virus and its high mortality have raised concerns.

**Methods:** To analyze evolution of outcomes, longitudinal clinical data and specimens were collected from five patients infected with H5N6 virus after admission. All patients received antiviral treatment either sequentially or in combination of oseltamivir with baloxavir. Severity of illness, and viral load in sputum, urine and blood, and cytokine levels in serum and sputum were serially analyzed.

**Results:** When delayed oseltamivir showed poor effects on high respiratory viral load, baloxavir was prescribed and viral load had a rapid reduction. All patients developed acute respiratory distress syndrome (ARDS) and sepsis within one week after disease onset, three patients died eventually. Nonsurvivors had more severe preexisting condition, extrapulmonary organ dysfunction and insufficient H5N6 virus-specific antibody response. Grouped by delta SOFA on the sample collection date, serum levels of IL-1α, IL-1β, IL-1RA, MIF, Mig, MIP-1α, IFN-γ, IL-12p40, IL-16, IL-18, IL-2Rα, IL-6, basic FGF, G-CSF, HGF, M-CSF, SCF were identified as indicator cytokines reflecting sepsis progression; and sputum levels of IL-18, IL-6, HGF, M-CSF were indicators of ARDS progression. Comparisons of cytokine levels before, during and after baloxavir treatment suggested that, baloxavir may also reduce a few indicator cytokines in sputum and serum that related to viral load and multi-organ dysfunction.

**Conclusions:** Baloxavir can effectively reduce viral load and few proinflammatory cytokines associated with deterioration. However, disease outcome is determined by severity of preexisting conditions and multi-organ dysfunction.

**Highlights:**

(1) Baloxavir potently decreased viral load in avian influenza H5N6 human infections.
(2) Preexisting conditions, extrapulmonary dysfunction and systemic inflammation determined prognosis of H5N6 patients.
(3) Indicator cytokines in sputum and serum reflecting ARDS and sepsis progression respectively, were identified in H5N6 patients.

## Introduction

Human infection with avian influenza A viruses can cause severe diseases, in which highly pathogenic avian influenza (HPAI) A viruses often induce fulminant pneumonia and multi-organ failure. The first outbreak of zoonotic avian influenza A viruses in human occurred in Hong Kong in 1997, with 6 deaths in 18 cases resulted from highly pathogenic avian influenza (HPAI) A subtype H5N1 virus infection [1]. As of January 2023, H5N1 infections have caused at least 868 human cases with 457 deaths worldwide (case fatality rate [CFR], 52.6%) [2]. Since 2013, continued reassortment of H5N1 viruses has generated multiple subtypes [3], in which novel HPAI H5N6 viruses of clade 2.3.4.4 have recently become the dominant H5 lineage circulating in China [4]. After the first case reported in 2014, most H5N6 human infections occurred in China, except for one case reported from Laos in 2021 [5]. As of 10 March 2023, totally 84 cases and 33 deaths of H5N6 human infection were reported globally (CFR, 39.3%) [6], in which 36 cases and 22 cases arose in 2021 and 2022, respectively, and such increase of human HPAI infection seems to continue [4, 7], posing potential public health threat.

Human H5N6 infections commonly initiated with fever and cough, which were difficult to distinguish from other respiratory infection at early stage [4, 8], and subsequently acute respiratory distress syndrome (ARDS), sepsis and multiple organ dysfunction (MOD) rapidly progressed [9, 10]. High viral load, prolonged viral shedding, and the elicited intense inflammatory response are central to the pathogenesis of HPAI human infections [11, 12], hence early antiviral treatment and timely immunomodulatory therapy play a crucial role in clinical management. However, the longitudinal viral and immunological associations with disease outcome remain unclear.

Neuraminidase inhibitors (NAI), usually oseltamivir monotherapy, has survival benefit for H5N1 and H7N9 patients especially when treatment initiated early in the clinical course [13, 14]. On the other hand, delayed NAI treatment proved to be an independent risk factors of prolonged viral shedding and NAI combination therapy could not reduce shedding duration in H7N9 patients [15]. Baloxavir marboxil is a novel small-molecule cap-dependent endonuclease inhibitor targets influenza polymerase acidic protein subunit, which has greater antiviral effects against influenza virus than oseltamivir, both in mouse models and uncomplicated influenza patients [16, 17]. Furthermore, baloxavir exhibits better antiviral effects and protection against lethal challenge compared with oseltamivir in lethal mouse models of avian influenza virus H7N9 [18] and H5N1 [19]. However, the effectiveness of baloxavir against HPAI human infection remains unknown.

Here, we reported five laboratory-confirmed human infection cases with avian H5N6 virus discovered in Guangdong province, China. All these patients received antiviral treatment using either sequentially or in combination of oseltamivir with baloxavir. We discovered indicator cytokines reflecting ARDS or sepsis progression and their correlations with viral load, virus-specific serological response and disease severity were analyzed. To evaluate potential effects of baloxavir, levels of indicator cytokines were compared before, during, and after treatment.

## Methods

### Design overview and participants

Five patients with laboratory-confirmed H5N6 virus infection between August 2021 and July 2022 in Guangdong Province, China, were included. As previously described, infection of avian influenza A(H5N6) virus was confirmed by metagenomic next-generation sequencing(mNGS) [4], reverse-transcriptase polymerase chain reaction (RT-PCR) assay [20] and serologic testing of hemagglutination inhibition assay and microneutralization assay [21]. Details of laboratory diagnosis are provided in the Supplementary Appendix.

The study was approved by ethical committees of The First Affiliated Hospital of Guangzhou Medical University (Ethics No. 2016-78). Written consents have been signed by the patients or their family members for collecting samples and clinical data for this study.

### Clinical data and sample collection

Medical records of five patients were collected and reviewed by a team of physicians who had clinical experience with avian influenza. Samples of different time points from each H5N6 patient were collected according to actual clinical situation, in which blood, throat swabs, sputum and bronchoalveolar lavage fluid (BALF) were accessible from all patients, while urine samples were able to obtain from patient 1, 2, 3 and 5.

### Identification and characterization of H5N6 virus from patients’ respiratory samples

Details of methods used to identify and characterize the H5N6 virus were provided in the Supplementary Appendix. Briefly, BALF samples from five patients were used for mNGS analysis; the eight gene fragments of H5N6 virus were amplified by conventional RT-PCR (Supplementary Table 1) and the PCR product was purified and sequenced. The H5N6 viral load was measured on sputum, throat swabs, peripheral blood and urine samples from the patients. Viral RNAs were extracted from heat inactivated clinical samples using the QIAamp Viral RNA Kit (Qiagen) in a biosafety II laboratory. Quantitative detection of viral nucleic acid in the above samples was then performed using the Detection Kit for Influenza A virus RNA (HUARUIAN BIOLOGY) and analyzed simultaneously according to the manufacturer’s instruction.

### Measurements of sequential organ failure assessment score

To quantify severity of organ dysfunction in H5N6 patients, admission and daily maximum Sequential Organ Failure Assessment (SOFA) [22, 23] was scored and reviewed by the team of experienced physicians as mentioned above. During assessment, some principles [24] were followed to ensure applicability and consistency, and details were provided in the Supplementary Appendix and Supplementary Table 2. Sepsis was diagnosed with SOFA score above two[22]. Delta SOFA score was calculated as the change of SOFA score from admission to the defined time point.

### Definitions

The time of disease onset was defined as the date of influenza-like illness (ILI) onset (fever and cough or sore throat) or other acute-onset signs or symptoms (e.g., fatigue and dyspnea) consistent with influenza illness, which was designated as day 0 after onset (D0). For dynamic measurements, the time of viral load, SOFA scores, cytokines and other laboratory indices were calculated from disease onset to the sample/data collection date. Viral shedding duration referred to the time from disease onset to the last positive result on RT-qPCR tests, in which threshold cycle values above 35 was defined as negative results. Definitions of pneumonia, ARDS and acute kidney injury are provided in the Supplementary Appendix.

### Antiviral treatment

NAI therapy (oseltamivir) was given according to local clinical practice. Baloxavir was administered when respiratory viral load remained above 5 log_10_ copies/mL after at least 4 days of NAI treatment, as 40 mg (for 40 kg to <80 kg bodyweight) or 80mg (for ≥80 kg bodyweight) enterally for every 3 days and terminated when influenza viral RNA was negative. Patient 1 and patient 4 received oseltamivir-baloxavir sequential treatment because of severe renal failure, and other patients received oseltamivir-baloxavir combined treatment (Table 1).

**Table 1.**
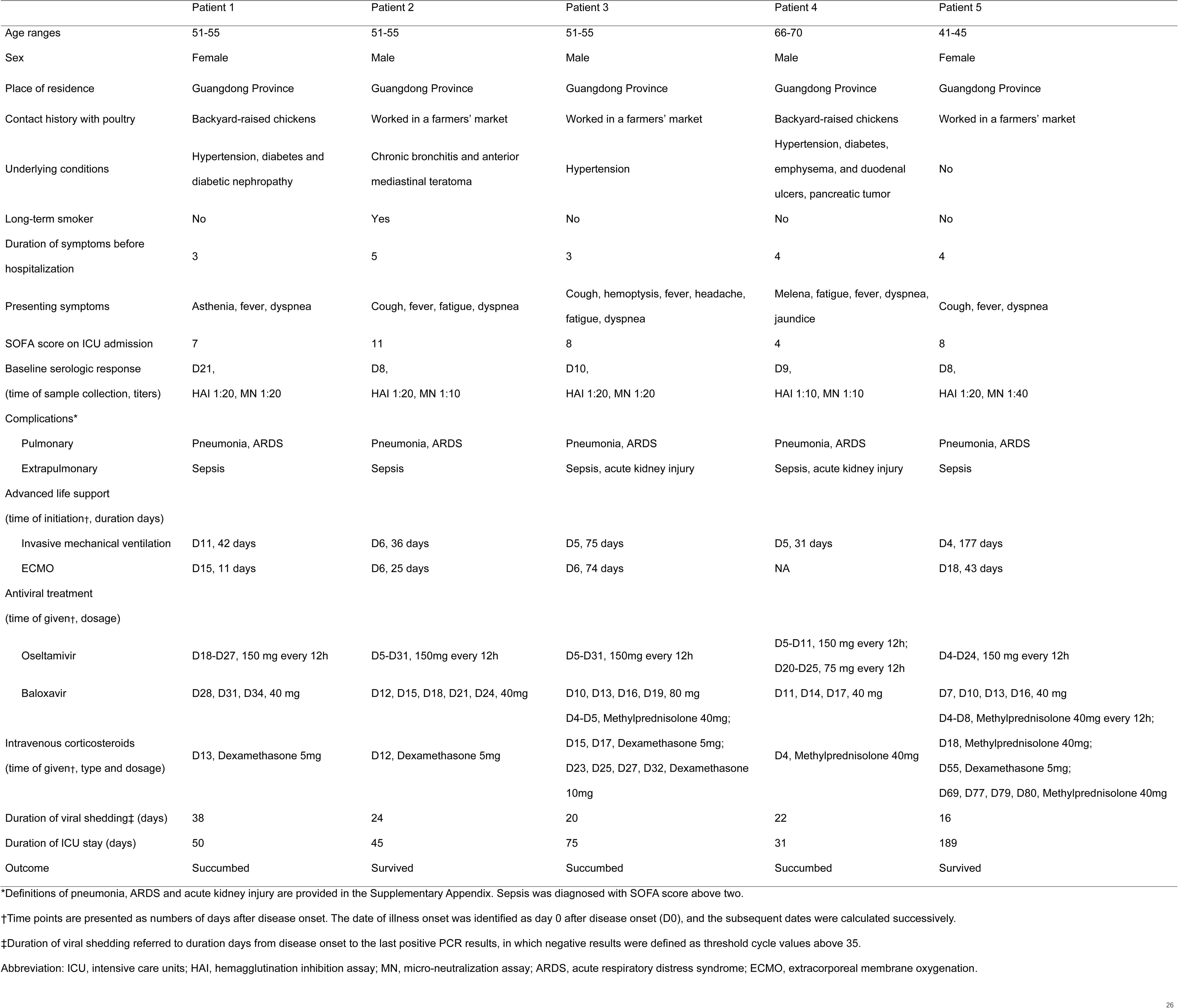
Epidemiologic and clinical characteristics of human infections with avian influenza A H5N6 virus in Guangdong Province, China from 2021 to 2022.

### Cytokine measurements

Sputum and serum samples of five patients were collected at multiple time points after hospitalization (Supplementary Table 3 and Supplementary Figure 1). Protein concentrations of 48 cytokines in sputum supernatants and serum were measured using the Bio-Plex Pro Human Cytokine Screening 48-Plex Panel on the Bio-Plex 200 Multiplex Testing System (Bio-Rad) following manufacturer instructions.

### Statistical analysis

Correlations between cytokines, viral load and clinical parameters were assessed by Spearman’s correlation tests. Serum cytokines at multiple time points from five patients were bulked and grouped according to delta SOFA at the corresponding time points, which included delta SOFA ≥ 0 or delta SOFA <0. Sputum cytokines were grouped by delta SOFA respiration sub-scores. Cytokine levels between groups were compared and significance was determined by Wilcoxon rank-sum tests. GraphPad Prism 9.0, R package corrplot (https://cran.r-project.org/web/packages/corrplot/) and ggpubr (https://cran.r-project.org/web/packages/ggpubr/) were used for analysis.

## Results

### Features of epidemiology, clinical manifestation and laboratory abnormalities in H5N6 virus infected patients

All five patients had contact history with poultry that had no signs of HPAI infection (Table 1). Patients with more severe underlying conditions (patient 1 and 4) had backyard-raised chickens, and the others with less severe chronic illness worked in the farmers’ market. Nonsurvivors had major comorbidities of hypertension (patient 1, and 4), diabetes (patient 1 and 4), and chronic obstructive pulmonary disease (COPD, patient 4). All patients initially presented with ILI symptoms and quickly developed into dyspnea and ARDS (PaO_2_/FiO_2_ <300) within one week, requiring invasive mechanical ventilation. With SOFA score above two, sepsis was diagnosed in five patients on ICU admission and as hypoxemia worsened, extracorporeal membrane oxygenation was administered in four patients (Figure 1A-1E and Supplementary Figure 1). As fulminant pneumonia developed, influenza was firstly suspected. However, with continuously searching for causative pathogens, avian influenza A (H5N6) virus wasn’t found in the patients’ respiratory tracts until mNGS on BALF (Supplementary Appendix). Phylogenetic analysis showed that the hemagglutinin (HA) genes of H5N6 virus derived from all five patients belonged to clade 2.3.4.4, whereas the neuraminidase genes (NA) belonged to Major clade among H5N6 serotypes [25] (Supplementary Figure 2). Serological analysis further confirmed the diagnosis (Figure 1F-1J).

**Figure 1.**
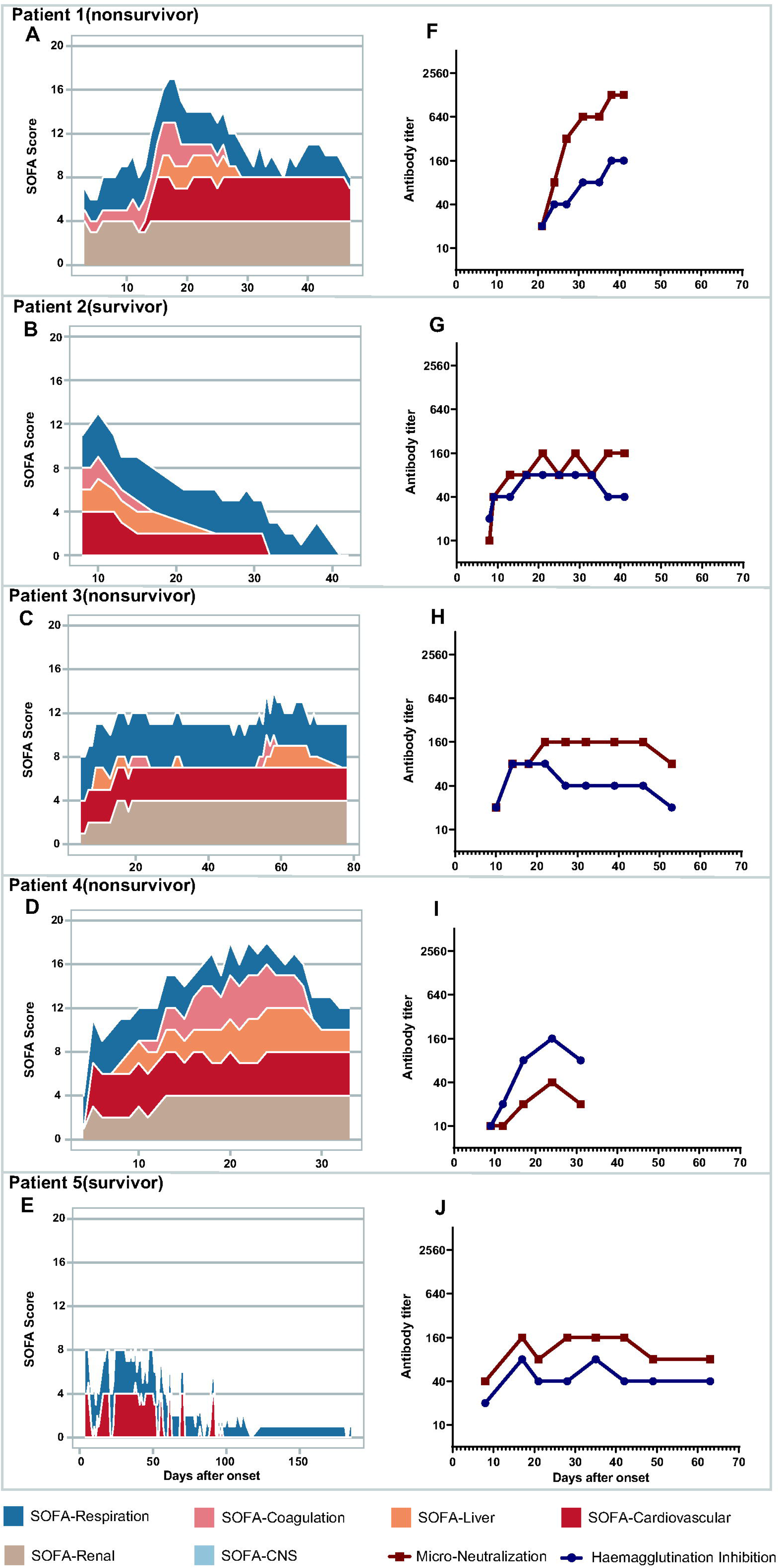
Figure 1. Dynamic change of SOFA scores and virus-specific serologic response in five H5N6 patients. A-E, Area charts show the temporal variations in daily maximum SOFA scores and sub-scores of five patients. F-J, Hemagglutinin inhibition (HAI) and microneutralization (MN) antibody titers of serum samples from patients at different time points after disease onset. HAI titers (blue circle) and neutralization titers (red square) are detected via recombinant (6+2) DG/H5N6-PR8 virus.

Three patients required renal replacement therapy because of preexisting diabetic nephropathy (patient 1) and acute kidney injury arose (patient 3 and 4). Intravenous corticosteroids were prescribed in all patients, in which patient 3 and 5 were given more than once. Characteristics of laboratory measurements in five patients were summarized in Supplementary Table 4. Serum creatinine in patient 1, 3 and 4 elevated on admission, suggesting renal dysfunction. In patient 1, 2, 3 and 5, bacterial infection was confirmed by positive sputum or blood culture results after at least 10 days of viral RNA detection. Three patients had bacteria cultured in sputum and multidrug-resistant bacteria emerged in patient 1 and patient 5. Staphylococcus epidermidis septicemia presented in patient 3. After at least 30 days in ICU, patient 1, 3 and 4 died from sepsis and multiple organ failure, and patient 2 and 5 survived.

Compared with survivors, nonsurvivors (patient 1, 3 and 4) had more dysfunctional organs involved, dominated by respiratory, cardiovascular and renal systems, in which SOFA scores gradually increased and endured until death (Figure 1A, 1C and 1D). Moreover, nonsurvivors had relatively low or delayed antibody response (Figure 1F, 1H and 1I).

### Antiviral effects of baloxavir treatment

High viral load (above 5 log_10_ copies/mL) was detected in sputum from all patients, while viral load in throat swabs was relatively low (Figure 2). For patient 2, viral RNA was detected in urine and duration of urine viral shedding was 25 days (Figure 2B). The median time from disease onset to oseltamivir prescription was 5 days for five patients (Table 1 and Supplementary Figure 1). Considering high sputum viral load and relatively poor effects of delayed NAI usage [12], baloxavir marboxil [17, 26] was prescribed. Median time from disease onset to baloxavir prescription was 11 days. In 4 patients (patient 1, 2, 3 and 4), median sputum viral load was above 5 log_10_ copies/mL after 5 days of double-dose oseltamivir, but was enormously reduced after baloxavir prescription, with over 200-fold reduction on average after twice baloxavir prescriptions (Figure 2A-2D). Median viral shedding duration of all patients was 22 days. Patient 5 who survived, had the shortest duration of viral shedding (16 days); the time from disease onset to baloxavir prescription was 7 days, which was the earliest initiation of baloxavir usage among five patients (Table 1 and Figure 2E). These results suggested that usage of baloxavir can potently and rapidly decrease the viral load in H5N6 patient even at the later stage of disease.

**Figure 2.**
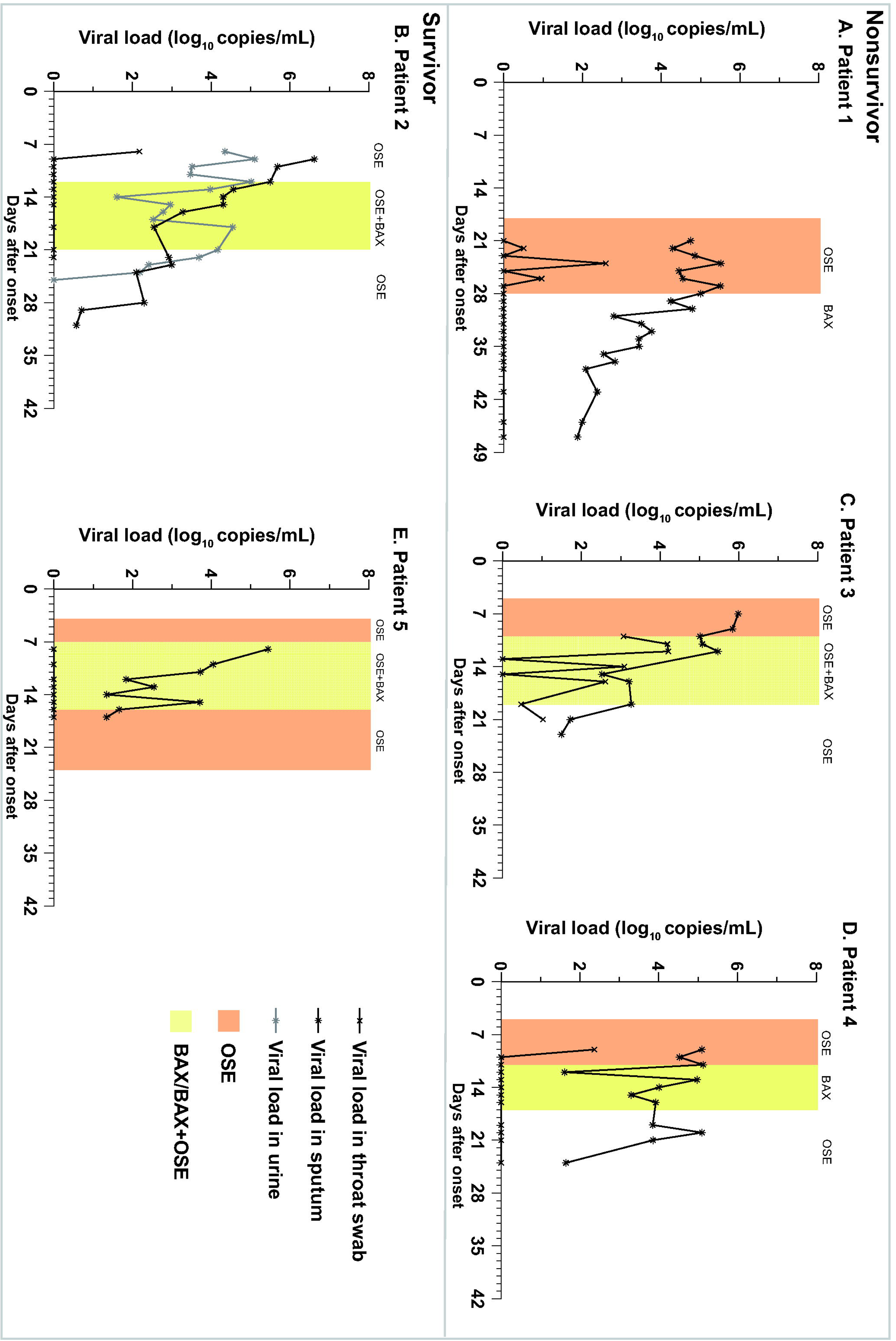
Dynamic change of viral load in five H5N6 patients. A-E, Viral dynamics and antiviral treatment. Respiratory viral load at different time points are marked by cross signs (throat swabs) and black asterisks (sputum). Viral load in urine from patient 2 are marked by gray asterisks. Filling color of light orange represents duration of oseltamivir prescription. Filling color of yellow represents duration of baloxavir prescription (one dose every 3 days), with or without oseltamivir. Abbreviation: OSE, oseltamivir; BAX, baloxavir.

Associations between viral dynamics with severity of hypoxemia and sepsis were analyzed respectively (Supplementary Figure 3). Statistically significant correlations between viral load with both PaO_2_/FiO_2_ ratio and SOFA scores were only found in two patients, indicating that H5N6 viral load in airways may not be a sole determinant for hypoxemia development and vital organ damage.

### Identification of indicator cytokines and the effect of baloxavir on their levels

For further investigation on immunological response, sputum and serum samples were collected from five patients to measure cytokine levels throughout their disease trajectories (Supplementary Figure 4 and 5). Strong and comparable correlation intensities were observed between sputum cytokines in survivors and nonsurvivors; however, serum cytokines were more pronounced in nonsurvivors than in survivors (Supplementary Figure 6).

Delta SOFA has been recommended in clinical practice [27] to dynamically evaluate severity of sepsis and organ dysfunction for its reliable association with mortality and low degrees of heterogeneity [24]. To discover cytokines with potential value of dynamic assessment, we first classified sampling timepoints as disease progression or regression by delta SOFA ≥ 0 or delta SOFA < 0 (Figure 3A). Cytokines with statistically significant differences between two kinds of timepoints were identified as indicator cytokines, which could reflect disease status of H5N6 patients. Among 48 cytokines, we found 4 cytokines in sputum and 18 cytokines in serum as indicators of ARDS progression and sepsis progression, respectively. Sputum IL-18, IL-6, HGF, M-CSF significantly elevated when ARDS deteriorated (Figure 3B). Serum IL-1α, IL-1β, IL-1RA, MIF, Mig, MIP-1α, IFN-γ, IL-12p40, IL-16, IL-18, IL-2Rα, IL-6, basic FGF, G-CSF, HGF, M-CSF, SCF significantly increased, Eotaxin significantly decreased when sepsis deteriorated (Figure 3C).

**Figure 3.**
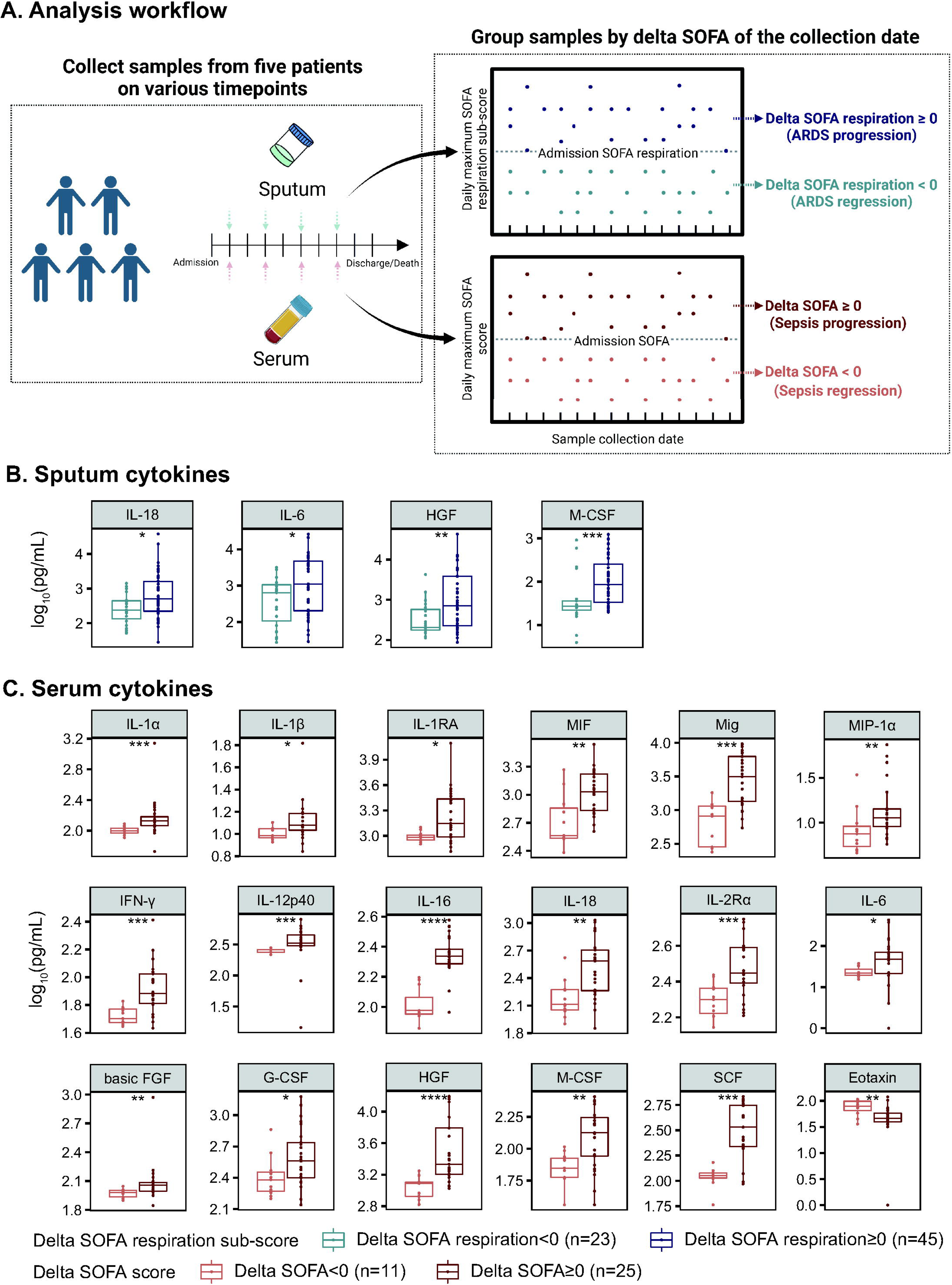
Comparison of cytokine levels during illness regression and progression to identify indicator cytokines. A, Analysis workflow of identifying indicator cytokines that potentially reflect ARDS or sepsis progression. Serum samples collected at multiple time points from five patients are grouped according to delta SOFA at the corresponding time points, sputum samples are grouped by delta SOFA respiration sub-scores. Delta SOFA is calculated by the change of SOFA scores/sub-scores from admission to the specific time point (detailed methods in Supplementary Appendix). At the sample collection date, delta SOFA ≥0 indicated disease progression, and delta SOFA < 0 indicated disease regression, respectively. And sputum/serum cytokine levels were compared between two kinds of disease status. B, Indicator cytokines in sputum. Box and jitter plots of sputum cytokine levels at ARDS regression (delta SOFA respiration sub-score on the collection date < 0, light blue, samples n=23) compared with those at ARDS progression (delta SOFA respiration sub-score on the collection date ≥ 0, dark blue, samples n=45), with raw values plotted on the log10 scale. C, Indicator cytokines in serum. Box and jitter plots of serum cytokine levels at sepsis regression (delta SOFA on the collection date < 0, orange, samples n=11) compared with those at sepsis progression (delta SOFA on the collection date ≥ 0, red, samples n=25), with raw values plotted on the log10 scale. Medians and interquartile ranges were presented. Dots represented individual data at different time points. Significance was determined by Wilcoxon rank-sum tests. *P* values: * *P*<0.05, ** *P*<0.01, *** *P*<0.001. Abbreviation: ARDS, acute respiratory distress syndrome.

Next, we analyzed correlations between indicator cytokines with days after disease onset (D.A.O), viral load, serum antibody titers and SOFA scores, respectively. Levels of indicator cytokines in sputum were all positively correlated with sputum viral load and SOFA scores, and HGF significantly decreased over D.A.O (Figure 4A). Except for IL-12p40 and G-CSF, all indicator cytokines in serum had positive correlation with SOFA scores, and the majority also had positive correlations with renal sub-scores (Figure 4B). Serum Mig and IL-18 were positively correlated with viral load, SOFA scores and four sub-scores, while IL-18 significantly decreased over D.A.O. Besides, lower titers of virus-neutralizing antibody correlated with higher IL-6 and IL-1β levels in serum. To evaluate impacts of baloxavir on indicator cytokine levels, sampling timepoints were divided as before/during/after baloxavir treatment, and the alterations in cytokine levels were compared. All 4 indicator cytokines in sputum significantly decreased after baloxavir treatment (Figure 4C). However, only Mig and IL-18 statistical-significantly decreased in serum indicator cytokines after baloxavir treatment (Figure 4D). The above results suggested that, indicator cytokines could reflect the disease status of H5N6 patients, correlating with viral load, antibody response, and MOD. Baloxavir predominantly reduced respiratory inflammatory response, but also could reduce a few systemic cytokines related to MOD, which may be partially attributed to its antiviral effect.

**Figure 4.**
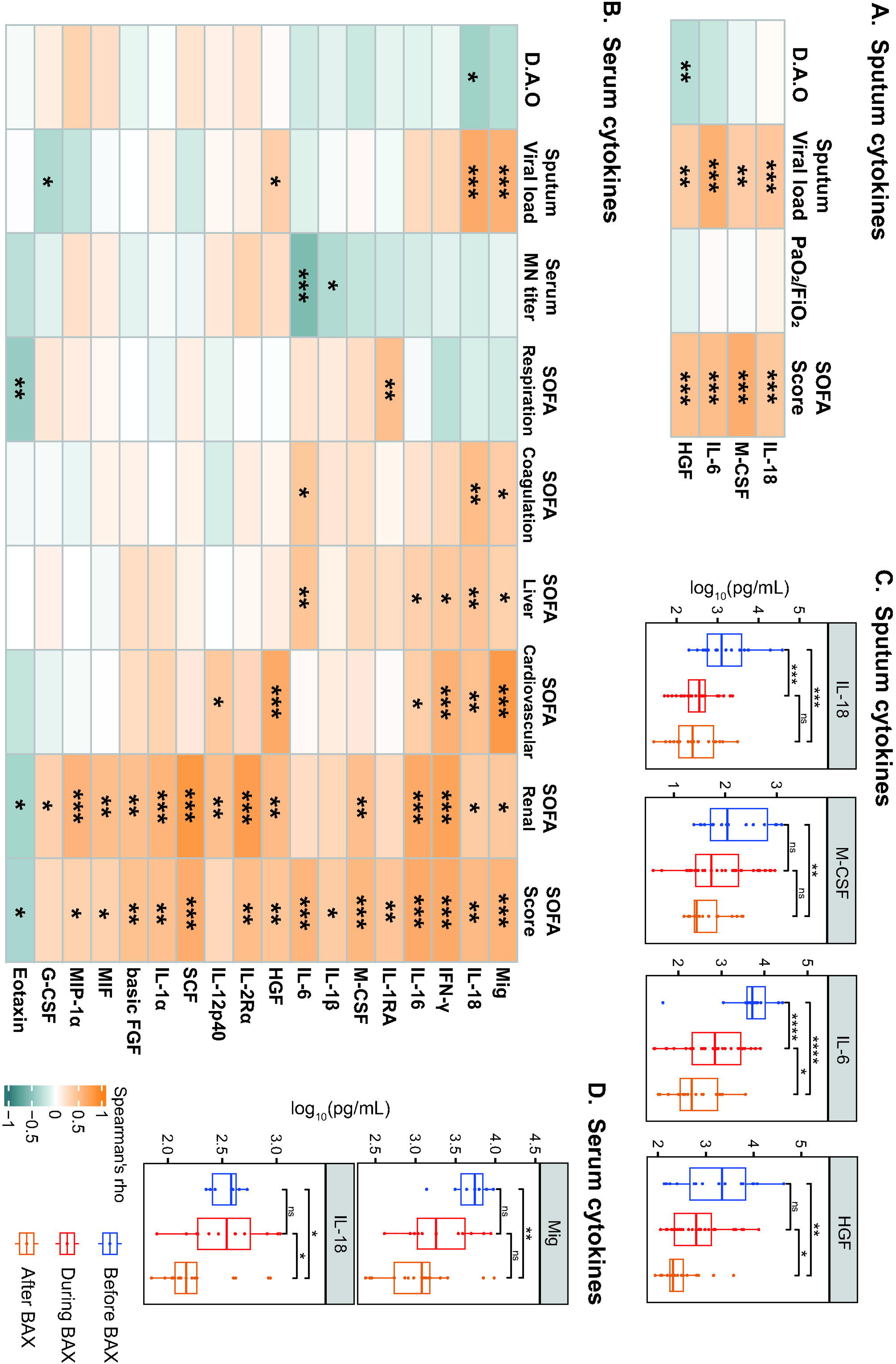
Correlation of indicator cytokine levels with viral load, clinical parameters and effect of baloxavir treatment. A, Heatmap showing correlation of sputum indicator cytokines with days after onset, sputum viral load, PaO_2_/FiO_2_ and SOFA scores. Significance was determined by Spearman’s correlation tests. B, Heatmap showing correlation of serum indicator cytokines with days after onset, sputum viral load, SOFA scores and sub-scores. Significance was determined by Spearman’s correlation tests. C, Effect of baloxavir treatment on sputum indicator cytokines. Box and jitter plots of indicator cytokine levels in sputum before, during and after baloxavir treatment (n=16, 31, 21, respectively). Significance was determined by Wilcoxon rank-sum tests. D, Effect of baloxavir treatment on serum indicator cytokines. Box and jitter plots of indicator cytokine levels in serum before, during and after baloxavir treatment (n=7, 12, 17, respectively). Significance was determined by Wilcoxon rank-sum tests. *P* values: * *P*<0.05, ** *P*<0.01, *** *P*<0.001, **** *P*<0.0001. Abbreviation: D.A.O, days after disease onset; BAX, baloxavir.

## Discussion

Limited by case numbers, previous summary of H5N6 human cases focused on comparisons with other influenza A virus infections [28] and lacked longitudinal data regarding disease pathogenesis. Our results revealed that, patients with underlying chronic illness was predisposed to H5N6 infection, which were also associated with fatality in influenza A(H1N1)pdm09 [29] and avian A(H7N9) [30] infections. H5N6 patients suffered from rapidly progressed and refractory sepsis, a life-threatening syndrome that mostly triggered by infection and characterized as systemic immune chaos with multi-organ failures [31]. Bacteria is frequently associated with sepsis development [32], yet viral sepsis has attracted attention for its high prevalence in COVID-19 [33]. The fact that sepsis developed in all H5N6 patients without evidence of bacterial infection within the first two weeks leads to a suspicion that it was triggered by high viral load, since influenza A virus was mostly detected in viral pneumonia-related sepsis [34]. Host immune response to influenza shares some common pathways with the response to bacteria [35, 36] might be an explanation. On the other hand, both seasonal and avian influenza infection were highly predisposed to bacterial infection, which is the leading cause of clinical deterioration [30, 36]. Positive cultures and elevated PCT revealed that, bacterial infections developed in all H5N6 patients after LRT viral load decreased and augmented organ dysfunction [32]. We proposed that H5N6 virus exacerbated underlying chronic illness and subsequently induced MOD, leading to poor outcomes.

As the incidence of HPAI infections in human rises [4], the search for more effective antiviral and immunomodulatory therapy becomes urgent. This is the first report of baloxavir for antiviral treatment in avian influenza human infection. Consistent with studies on severe influenza patients [26], baloxavir-NAI combination exhibited potent antiviral effect without outcome advantage. Our results showed that baloxavir could potentially decrease lung inflammation in H5N6 human infections, compatible with the efficacy in H5N1 murine models [19], yet had limited influence on serum cytokines. Inadequate virus-specific serological response and hypercytokinemia may account for such unsatisfactory outcome, raising the possibility of using antibody-based treatment and immune modulators. In our data, the patient who took baloxavir within one week after illness onset had the shortest duration of viral shedding and survived, even with intravenous corticosteroids prescription that could prolong viral clearance [37], suggesting the coordination of potent antivirals and immune modulators has potential therapeutic benefit for extreme circumstances of avian influenza human infections.

While most cytokines in respiratory tract decreased with time and viral load, systemic inflammation dominated in H5N6 patients. Among the serum indicator cytokine profiles we discovered, IL-6, M-CSF and HGF have been previously identified as markers reflecting organ damage in severe COVID-19 patients [38]. Notably, taking together both our results and previous comparisons of serum cytokine profiles between H5N6, H7N9 and H1N1 patients [28], we speculate that serum IL-2Rα, IL-12p40 and M-CSF play pivotal roles in H5N6-induced sepsis and could be biomarkers of critical cases. Increased concentration of serum IL-2Rα is the result of T-cell activation [39], while IL-12p40 mainly acts as chemoattractant and M-CSF is colony-stimulating factor both for macrophages. Hypercytokinemia in H5N6 patients suggests the potential role of anti-cytokine antibodies and cytokine modulators [40], such as IL-6 receptor blockade and Janus kinase inhibitors [41].

It is also interesting to note that H5N6 virus RNA was detected in urine from one survivor (patient 2), while no evidence of viremia or renal dysfunction presented. This is the first report on urine viral shedding in H5N6 patients, which has been previously found in human infected with influenza A(H7N9) virus [42]. No association between urine viral RNA detection and renal dysfunction could be possibly accounted for hematogenous dissemination through infected lymphocytes or macrophages [42, 43]. There are limitations in this research. Case numbers were constrained by the sporadic occurrence of H5N6 human infections. As a retrospective summary of five cases, influence of systemic corticosteroids on secondary bacterial infection and outcome was unclear, which could also confound the cytokine profile analysis, remaining an ongoing challenge. While our case study provides some insights on antiviral treatment for HPAI human infections, the variability in patients and approach makes it difficult to draw string conclusions and therefore highlights the need for a prospective hypothesis driven study. It is interesting to investigate whether the initiation of baloxavir prescription as early as possible once patients show influenza-like symptoms in an HPAI endemic area, and the application of immunomodulators when hypercytokinemia develops, could lower the CFR of HPAI human infections.

## Conclusion

In summary, our study illustrates that for human infection with H5N6 virus, baloxavir can effectively reduce viral load and prevent pulmonary deterioration even at the later stage of disease. However, preexisting conditions, extrapulmonary dysfunction and systemic inflammation are potential determinants of the disease outcomes.

## Notes

## Data Availability

All data produced in the present study are available upon reasonable request to the corresponding authors.

## Acknowledgements

We thank Vision Medicals Laboratory and Guangzhou Huayin Medical Laboratory Center Co., Ltd. for their technical support on metagenomic next-generation sequencing. Also, we would like to thank Dr. Faqiang He, Dr. Zhipeng Li, Dr. Ruiyu Lu, Dr. Hanmian Liu, Dr. Yongli Wu, Dr. Zhihui Zhuang, and the ICU teams for taking care of the patients.

## Disclaimer

The funders had no role in the study design, results, interpretation, or decision to submit the manuscript for publication.

