## Supplementary Appendix for "Baloxavir marboxil use for critical human infection of avian influenza A H5N6 virus"

### Table of Contents


1. Methods of metagenomic next-generation sequencing (mNGS) and analysis
2. Methods of conventional reverse-transcription polymerase chain reaction
3. Methods of phylogenetic analysis of H5N6 viruses
4. Methods of hemagglutination inhibition assay and microneutralization assay
5. Methods of sequential organ failure assessment score measurements
6. Definitions of pneumonia, acute respiratory distress syndrome (ARDS) and acute kidney injury
7. Supplementary Table 1: List of primer sequences
8. Supplementary Table 2: Sequential organ failure assessment (SOFA) score
9. Supplementary Table 3: Collection time of serum and sputum samples for cytokine evaluation
10. Supplementary Table 4: Characteristics of laboratory measurements in five patients with avian influenza A H5N6 virus infection
11. Supplementary Figure 1: Key events timeline of five patients infected with influenza A(H5N6) virus in Guangdong Province, China, 2021-2022
12. Supplementary Figure 2: Phylogenetic trees of HA and NA genes of influenza A (H5N6) viruses in five patients in Guangdong Province, 2021 to 2022
13. Supplementary Figure 3: Correlations of $PaO_2/FiO_2$ ratio and SOFA scores with sputum viral load in five H5N6 patients
14. Supplementary Figure 4: Dynamic change of sputum cytokine concentrations in H5N6 patients over time
15. Supplementary Figure 5: Dynamic change of serum cytokine concentrations in H5N6 patients over time
16. Supplementary Figure 6: Correlation matrix across all time points of 48 cytokines in sputum/serum from five H5N6 patients with different outcomes
17. References
18. Legends of Supplementary Figures

**Methods of metagenomic next-generation sequencing(mNGS) and analysis**

RNA was extracted from BALF samples of five patients with the use of the QIAamp Viral RNA Kit (Qiagen), according to the manufacturer's instructions. Total RNA was then reverse transcribed and amplified using the VAHTS Universal V6 RNA-seq Library Prep Kit (Vazyme) for library construction as described by Zhang and colleagues [1]. Library concentration was assessed by Qubit dsDNA HS Assay kit and library pools were then loaded onto an Illumina Nextseq 550 sequencer to generate approximately 20 million reads for each library [2].

For library of each patient, low quality reads, adaptor sequences, non-complex reads and duplicated reads were removed using the Fastp (v0.22.0). The remaining reads were mapped against reference genome hg38 using BWA. After the human reads were removed, host-filtered reads were aligned to microorganism reference database with Bowtie2 (v2.3.5.1), which included influenza A(H5N6) virus genome (Combination of following strains: A/Cygnuscolum bianus/Hubei/52/2020 (MW505397), A/chicken/Ganzhou/ GZ157/2016 (KY415942), A/duck/Fujian/11.3_FZHX1132-O/2016 (MW107444), A/duck/Mongolia/729/2019 (MT020250), A/Goose/Guangdong/1/96 (NC_ 007361). Finally, aligned reads were extracted using samtools (v1.13) for genome sequence assembly. Aligned reads of these three samples were assembled in order to fetch full-length individual genome sequence using SPAdes (v3.15.3), with a single parameter named "rnaviral". Genome sequences of A/Huizhou/1/2021(H5N6), which obtained from patient 1 were deposited in the GenBank database of NCBI (Accession No.: OK284448-OK284453).

**Methods of conventional reverse-transcription polymerase chain reaction**

First-strand cDNA was synthesized using PrimeScript™ II 1$^{st}$ Strand cDNA Synthesis Kit (TaKaRa) using specific primer (10 μM, 5′-AGCAAAAGCAGG -3′)

primers, according to the vendor's protocol. The cDNA was subsequently stored at -20 °C.

Primers used in the amplification of eight gene fragments of H5N6 virus were shown in Supplementary Table 1. The PCR were performed in a reaction mixture consisting of 2.0 μL of each primer (10 μM of upstream and downstream), 25.0 μL of PrimeSTAR Max Premix (TaKaRa), 2.0 μL cDNA and 19 μL RNAase-free water. The thermal conditions were as follows: initial denaturation at 95 °C for 4 min; 35 cycles at 98 °C for 10 s, 55 °C for 15 s, and 72 °C for 50 s; and a final extension at 72 °C for 4 min. The PCR product was purified and for sequencing. Genome sequences of patient 2 (A/Dongguan/2/2021(H5N6)) were deposited in the GenBank database of NCBI (Accession No.: OL519551-OL519558).

**Methods of phylogenetic analysis of H5N6 viruses**

Phylogenetic analysis was performed by Maximum Likelihood based on 4846 H5(H5Nx) and 2745 N6(HxN6) viruses isolated from animals including mammals and avian species available in Global Initiative on Sharing All Influenza Data (GISAID) between 2011 and 2020, arbitrarily chosen 1190 H5 strains and 1190 N6 strains were analyzed. Multiple sequence alignment was performed using MAFFT (v7.487). A maximum likelihood tree using the HA genes rooted to A/goose/Guangdong/1/96(NC_007361) and NA genes was constructed for IQtree with 1000 replicate. Horizontal distances are proportional to the genetic distance.

**Methods of hemagglutination inhibition assay and microneutralization assay**

Hemagglutination inhibition (HI) assay and microneutralization (MN) assay were performed on the patients' sera to reconfirm H5N6 infection, according the protocol from World Health Organization [3]. A (6+2) reassortant viruse (DG/H5N6-PR8), containing the HA and NA gene of A/Dongguan/2/2021 (H5N6) in the gene background of A/Puerto Rico/8/1934 (PR8) virus was rescued by reverse genetics and used in detection.

For HI assay, sera samples were heat inactivated at 56 °C for 30 min and two-fold serially diluted (initial dilution 1:10) in 25 µL of PBS in V bottom plates, then an equal volume of 4 hemagglutinin (HA) units of corresponding reassortant virus were added and incubated for 30 min at room temperature. Fifty microliters of 1% chicken red blood cells (RBCs) suspension were added to the wells and incubated for 30 min. Reciprocal of serum dilution in last well showing complete hemagglutination inhibition was considered as HAI titer. The detection limit was a titer of 1:10.

For MN assay, sera samples were heat inactivated at 56 °C for 30 minutes and diluted 10 times with viral diluent (DMEM containing 1% BSA, 1% Penicillin-Streptomycin Solution and 2 µg/mL of TPCK-treated trypsin) and further two-fold dilution were prepared in 96-well cell culture plates with final volume of 50 µL in each well. Sera samples were then incubated with 100 $TCID_{50}$ of reassortant virus for 1 hour at 37 °C and 5% $CO_2$. 100 uL of 3 x $10^5$ cells/mL MDCK cells was added to each well and the plate was incubated for 22 hours at 37 °C and 5% $CO_2$. Plates were washed with PBS and fixed with cold 80% acetone for 20 minutes. Viral protein was detected by ELISA with anti-influenza A-NP antibody. The neutralizing antibody titer was defined as the reciprocal of the highest serum dilution that neutralized the virus in MDCK cell cultures. The minimum detection limit of this assay was a titer of 1:10.

**Methods of sequential organ failure assessment score measurements**

SOFA scores were measured according to the criteria provided in Supplementary Table 2. To ensure applicability and consistency, some principles[4] were followed during assessment: (1) The central nervous system sub-score would be carried over from the last score before intubation throughout the duration of sedative medication administration. (2) For patients undergoing renal replacement therapy, a renal sub-score of four would be applied. (3) Missing data at a single time point would be replaced by the mean of the preceding and immediately succeeding values, while missing data in two consecutive time points leading to the value be treated as a missing data point.

### Definitions of pneumonia, acute respiratory distress syndrome (ARDS) and acute kidney injury

Pneumonia was diagnosed as acute onset of cough and at least one of new focal chest signs, fever > 4 days or dyspnea/tachypnea, accompanied by chest radiograph findings of ground-glass opacity [5].

Acute respiratory distress syndrome (ARDS) was diagnosed based on degree of hypoxemia ($PaO_2/FiO_2 \leq 300$ mmHg with PEEP ≥ 5 cmH2O), accompanied with bilateral opacities on chest X-ray, which could not be fully explained by cardiac failure or fluid overload [6].

According to the clinical practice guidelines from KDIGO (The Kidney Disease: Improving Global Outcomes) [7], acute kidney injury was defined with any of the followings: increase in serum creatinine (serum CRE) by ≥ 0.3 mg/dL (≥ 26.5 μmol/L) within 48 hours; or increase in serum CRE to ≥ 1.5 times baseline, which is known or presumed to have occurred within the prior 7 days; or urine volume < 0.5 mL/kg/h for 6 hours.

**Supplementary Table 1. List of primer sequences**

| Primer Pair | Sequences |
|---|---|
| HA-H5 | F-5’-AGCAAAAAGCAAAAGCAGGGGTTCACTCTGTC -3’ |
| | R-5’-AGTAGAAACAAGGGTGTTTTTAATTAC-3’ |
| NA-N6 | F-5’-AGCAAAAGCAGGGTGAAAATGAATC-3’ |
| | R-5’-AGTAGAAACAAGGGTGTTTTTTC-3’ |
| H5N6-M | F-5’-GCAAAAGCAGGTAGATATTGAAAGATGAGTCT-3’ |
| | R-5’-AGTAGAAACAAGGTAGTTTTTTACTCCAGCTCTAT-3’ |
| H5N6-NP | F-5’-AGCAAAAGCAGGGTAGATAATCACTCACTGAGTGAC-3’ |
| | R-5’-AGTAGAAACAAGGGTATTTTTCTTTAATTGTCAT-3’ |
| H5N6-NS | F-5’-AGCAAAAGCAGGGTGACAAAGACATAATGGATTC-3’ |
| | R-5’-AGTAGAAACAAGGGTGTTTTTTATCATTAAATAAGCTG-3’ |
| H5N6-PB1 | F-5’-AGCAAAAGCAGGCAAACCATTTGAATGGATGTCAA-3’ |
| | R-5’-ACTATTTTTGCCGTCTGAGCTCTTCAATG-3’ |
| H5N6-PB2 | F-5’-AGCAAAAGCAGGTCAAATATATTCAATATG-3’ |
| | R-5’-AGTAGAAACAAGGTCGTTTTTAAACAATT-3’ |
| H5N6-PA | F-5’-AGCGAAAGCAGGTACTGATCCGAAATGGA-3’ |
| | R-5’-GGTAGAAACAAGGTACTTTTTTGGACAGTATG-3’ |

Abbreviations: F, forward; R, reverse.

**Supplementary Table 2. Sequential Organ Failure Assessment (SOFA) Score***

| | |
|---|---|
| **Respiration system** | |
| $PaO_2/FiO_2$, mmHg | SOFA respiration sub-score |
| ≥ 400 | 0 |
| < 400 | 1 |
| < 300 | 2 |
| < 200 with respiratory support | 3 |
| < 100 with respiratory support | 4 |
| **Coagulation** | |
| Platelets, ×$10^3$/μL | SOFA coagulation sub-score |
| ≥ 150 | 0 |
| < 150 | 1 |
| < 100 | 2 |
| < 50 | 3 |
| < 20 | 4 |
| **Liver** | |
| Bilirubin, μmol/L | SOFA liver sub-score |
| < 20 | 0 |
| 20 - 32 | 1 |
| 33 - 101 | 2 |
| 102 - 204 | 3 |
| > 204 | 4 |
| **Cardiovascular system** | |
| Mean arterial pressure (MAP) or required vasopressors | SOFA cardiovascular sub-score |
| MAP ≥ 70 mmHg | 0 |
| MAP < 70 mmHg | 1 |
| Dopamine < 5 μg/kg/min or dobutamine (any dose) | 2 |
| Dopamine 5.1 - 15 μg/kg/min or epinephrine ≤ 0.1μg/kg/min or norepinephrine ≤ 0.1 μg/kg/min | 3 |
| Dopamine > 15 μg/kg/min or epinephrine > 0.1 μg/kg/min or norepinephrine > 0.1 μg/kg/min | 4 |
| **Central nervous system (CNS)** | |
| Glasgow Coma Scale score | SOFA CNS sub-score |
| 15 | 0 |
| 13-14 | 1 |
| 10-12 | 2 |
| 6-9 | 3 |
| < 6 | 4 |
| **Renal** | |
| Serum creatinine, μmol/L; urine output | SOFA renal sub-score |
| < 110 | 0 |
| 110 - 170 | 1 |
| 171 - 299 | 2 |
| 300 - 440 (or urine output < 500 mL/d) | 3 |
| > 440 (or urine output < 200 mL/d) | 4 |
| **SOFA score** | Sum of each sub-score |
| **Admission SOFA score†** | Sum of the most severe value for each sub-score in the 24 h preceding admission to ICU |
| **Daily maximum SOFA score†** | Sum of the most severe values of each sub-score for each 24h assessment |
| **Delta SOFA score†** | Change of SOFA score from admission to a defined time point (For example, delta SOFA score on D3 = maximum SOFA score on D3 - admission SOFA score) |

*Derived from Singer et al.[8] and Lambden et al.[4]

†Definitions of admission SOFA, daily maximum SOFA and delta SOFA are also applicable to each SOFA sub-score.

Abbreviation: $PaO_2$, partial pressure of oxygen in arterial blood; $FiO_2$, fraction of inspired oxygen.

**Supplementary Table 3. Collection time of serum and sputum samples for cytokine evaluation**

| Sample type | Patients | Sample numbers | Time of collection (days after disease onset) |
|---|---|---|---|
| | Patient 1 | 7 | 21,24,27,31,35,38,41 |
| | Patient 2 | 10 | 8,9,13,17,21,25,29,33,37,41 |
| Serum | Patient 3 | 9 | 10,14,18,22,27,32,39,46,53 |
| | Patient 4 | 5 | 9,12,17,24,31 |
| | Patient 5 | 5 | 8,17,28,42,63 |
| | Patient 1 | 21 | 21,22,23,24,25,26,27,28,29,30,31,32,33,34,35,36,37,38,41,45,47 |
| | Patient 2 | 17 | 9,10,12,13,14,15,16,18,22,23,24,28,29,31,35,37,39 |
| Sputum | Patient 3 | 10 | 7,8,10,12,16,18,19,21,23,36 |
| | Patient 4 | 10 | 9,11,12,13,14,16,19,20,22,24 |
| | Patient 5 | 10 | 8,10,11,12,17,22,28,36,42,63 |

**Supplementary Table 4. Characteristics of laboratory measurements in five patients with avian influenza A H5N6 virus infection**

| Characteristics | Patient 1 | Patient 2 | Patient 3 | Patient 4 | Patient 5 | Abnormal range |
|---|---|---|---|---|---|---|
| Blood test results (results on admission, range during hospitalization) | | | | | | |
| White blood cells, $\times 10^9$/L | 4.60, 2.00-31.40 | 5.70, 4.99-15.72 | 8.59, 4.69-30.88 | 8.01, 3.37-34.42 | 3.80, 3.80-25.80 | <4.00, leukopenia; >10.00, leukocytosis |
| Lymphocytes, $\times 10^9$/L | 0.39, 0.11-1.75 | 0.52, 0.52-3.66 | 0.64, 0.45-2.62 | 0.66, 0.27-2.13 | 0.45, 0.18-3.24 | <1.00, lymphopenia; >4.00, lymphocytosis |
| Neutrophils, $\times 10^9$/L | 3.58, 1.76-30.68 | 5.10, 3.46-11.40 | 7.64, 2.88-27.61 | 7.22, 2.59-29.22 | 3.30, 2.73-22.88 | <1.50, neutropenia; >8.50, neutrophilia |
| Platelets, $\times 10^9$/L | 145.00, 14.00-371.00 | 123.00, 72.00-354.00 | 192.00, 88.00-376.00 | 251.00, 3.00-264.00 | 200.00, 131.00-717.00 | <150.00, thrombocytopenia; >450.00, thrombocytosis |
| Hemoglobin, g/L | 49.00, 49.00-128.00 | 150.00, 78.00-150.00 | 318.00, 72.00-318.00 | 85.00, 70.00-112.00 | 106.00, 74.00-134.00 | <132, low hemoglobin count in men; <116, low hemoglobin count in women |
| PCT, ng/mL | 0.32, 0.24-16.65 | 0.34, 0.05-1.16 | 0.18, 0.18-2.24 | 16.01, 4.09-98.89 | 0.64, 0.01-0.83 | >0.5, elevated PCT |
| ALT, U/L | 17.00, 1.00-58.00 | 365.00, 14.90-365.00 | 93.20, 2.40-109.30 | 54.50, 8.00-54.50 | 32.00, 9.00-89.00 | >50.00, elevated ALT |
| AST, U/L | 11.00, 9.00-568.00 | 173.00, 22.60-173.00 | 59.40, 13.00-462.50 | 47.90, 33.60-238.50 | 98.00, 10.00-248.00 | >50.00, elevated AST |
| Total bilirubin, μmol/L | 5.70, 5.70-58.90 | 74.10, 11.60-124.10 | 12.30, 4.60-71.50 | 9.10, 8.45-316.77 | 11.50, 2.10-23.70 | >17.00, hyperbilirubinemia |
| LDH, U/L | 309.00, 226.00-1509.00 | 728.10, 196.00-1112.00 | 400.20, 219.60-1840.90 | 242.95, 196.88-764.00 | 798.00, 204.00-1210.00 | >250.00, elevated LDH |
| CK, U/L | 170.00, 19.00-887.00 | 259.20, 73.00-554.00 | 435.00, 20.00-435.00 | 19.80, 19.80-898.10 | 134.00, 4.00-702.00 | >200.00, elevated CK |
| Serum CRE, μmol/L | 700.00, 40.00-700.00 | 58.00, 26.00-74.00 | 123.00, 108.00-548.00 | 164.20, 74.50-336.90 | 55.00, 8.00-64.00 | >120.00, elevated CRE |
| Bacterial infection (time of collection†, bacteria) | | | | | | |
| Positive sputum culture | D27, Enterobacter cloacae and Klebsiella pneumoniae (ESBLs+); D44, Pseudomonas aeruginosa | D22, Stenotrophomonas maltophilia | None | None | D47, D49, D52, Sphingomonas paucimobilis; D161, D163, Pseudomonas aeruginosa (CRPA) | |
| Positive blood culture | None | None | D16, Staphylococcus epidermidis | None | None | |

†Time points are presented as numbers of days after disease onset. The date of disease onset was identified as day 0 after disease onset (D0), and the subsequent dates were calculated successively.

Abbreviation: PCT, procalcitonin; ALT, alanine aminotransferase; AST, aspartate aminotransferase; LDH, lactate dehydrogenase; CK, creatine kinase; CRE, creatinine; ESBLs, extended-spectrum beta-lactamases; CRPA, carbapenem-resistant Pseudomonas aeruginosa.

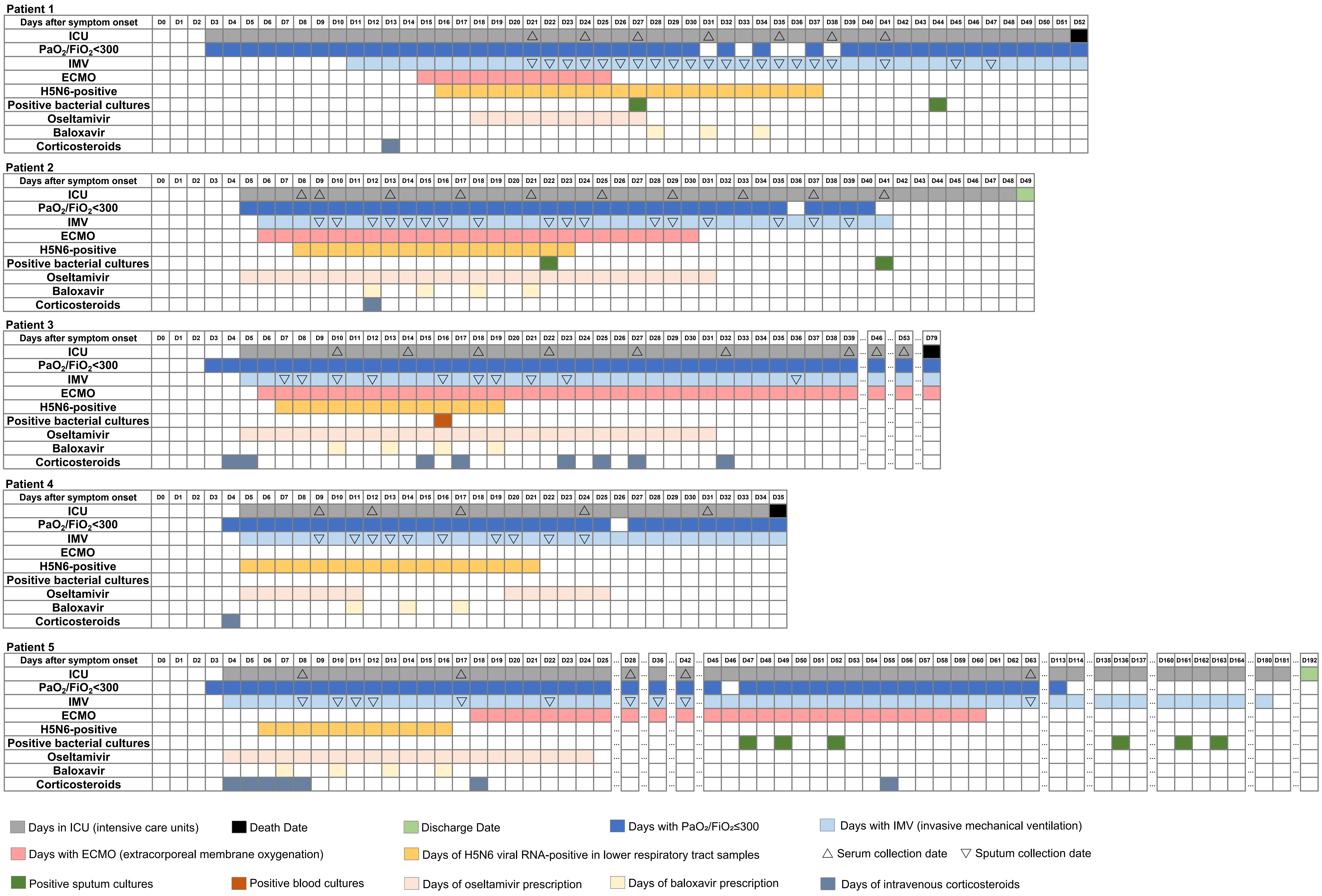


**Supplementary Figure 1. Key events timeline of five patients infected with influenza A(H5N6) virus in Guangdong Province, China, 2021-2022**

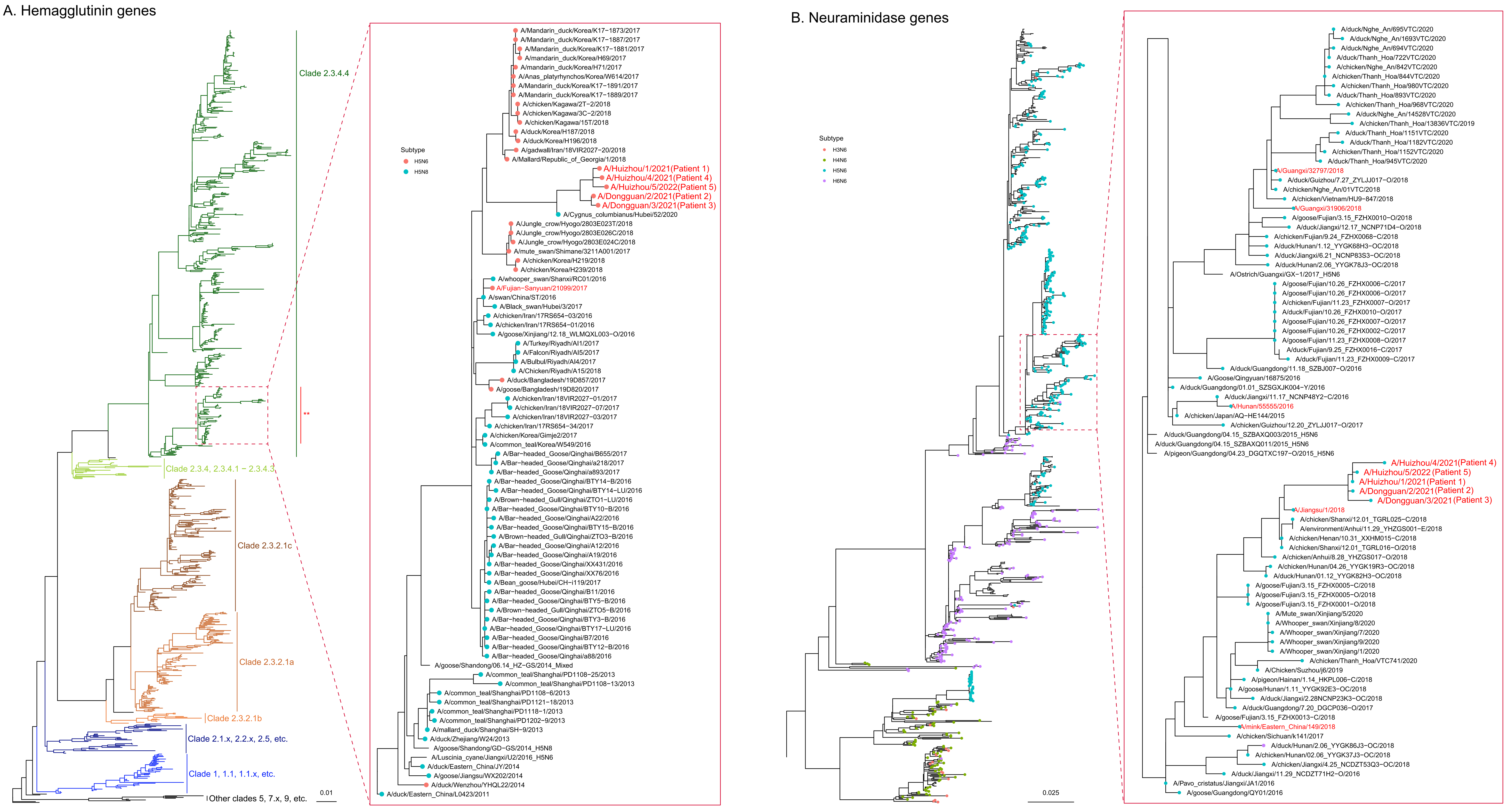


**Supplementary Figure 2. Phylogenetic trees of HA and NA genes of influenza A (H5N6) viruses in five patients in Guangdong Province, 2021 to 2022**

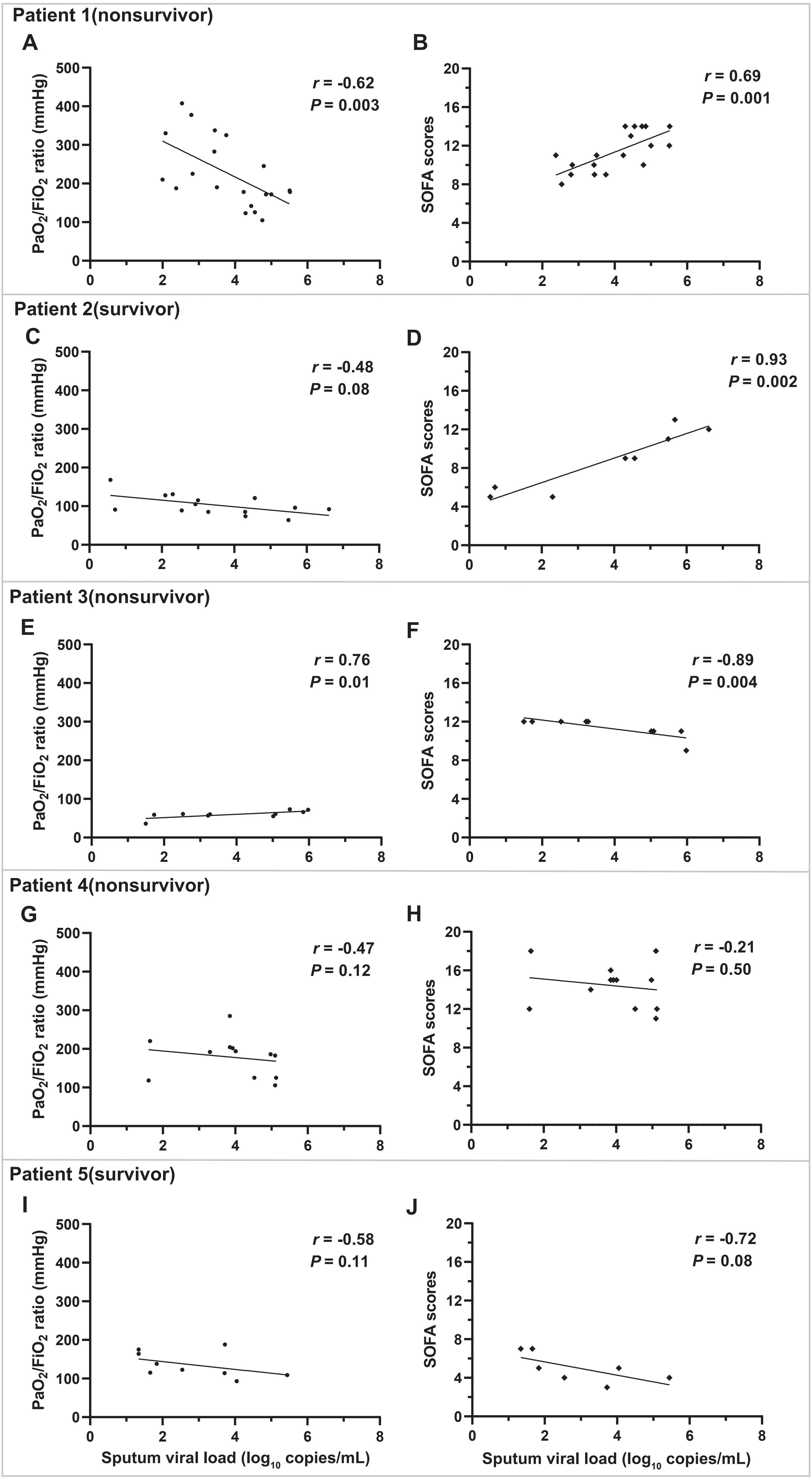


**Supplementary Figure 3. Correlations of $PaO_2/FiO_2$ ratio and SOFA scores with sputum viral load in five H5N6 patients**

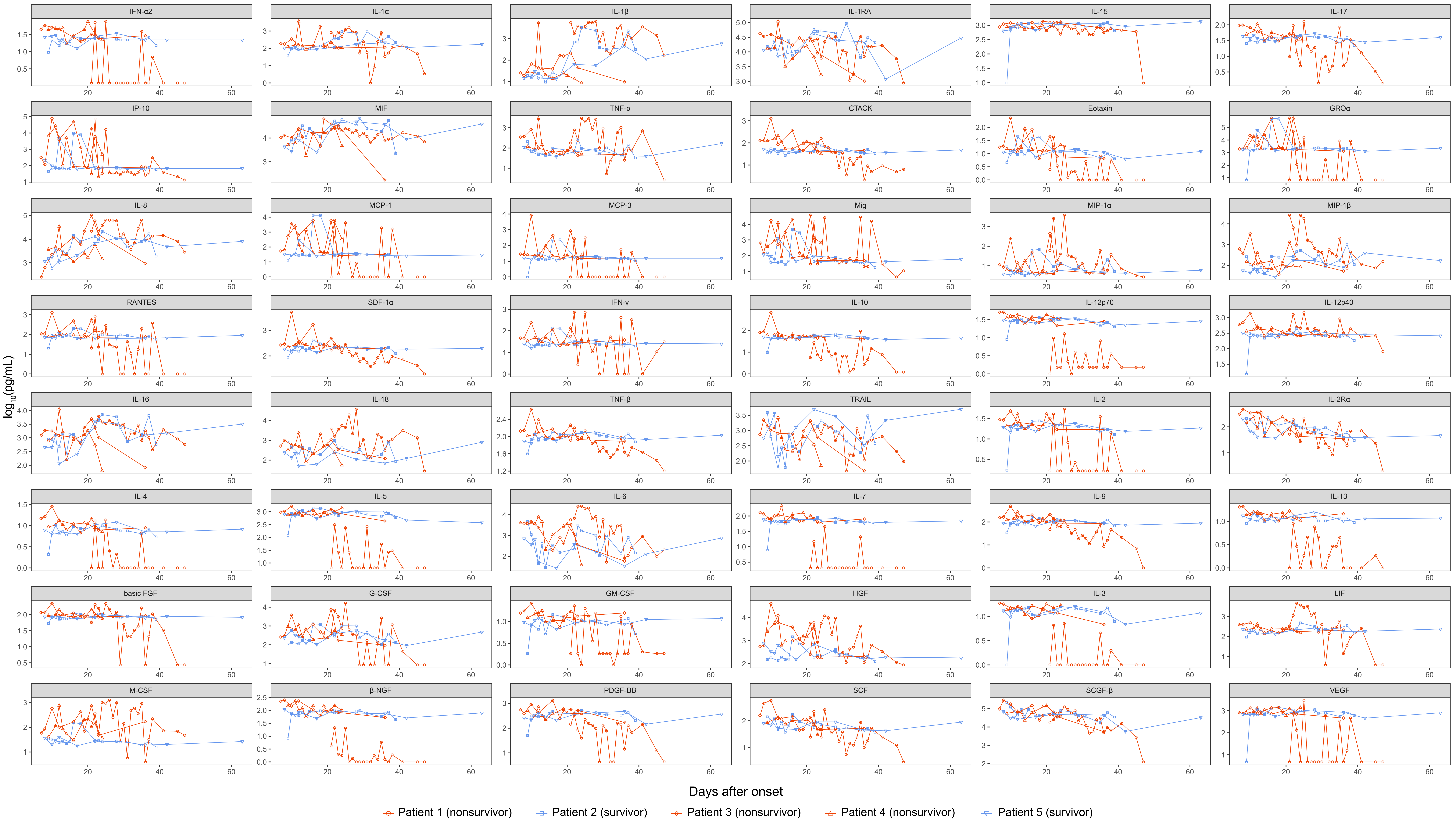


**Supplementary Figure 4. Dynamic change of sputum cytokine concentrations in H5N6 patients over time**

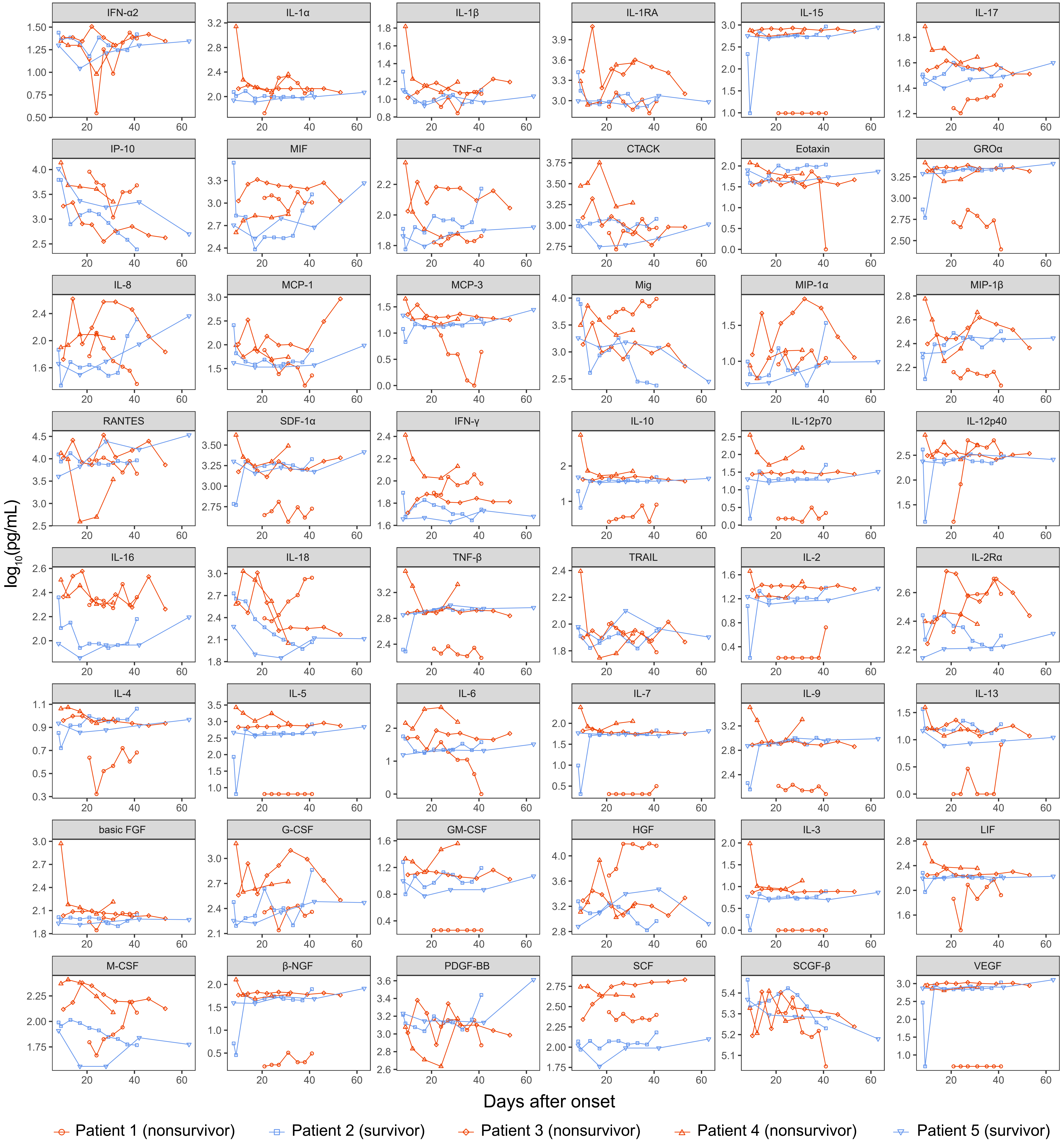


**Supplementary Figure 5. Dynamic change of serum cytokine concentrations in H5N6 patients over time**

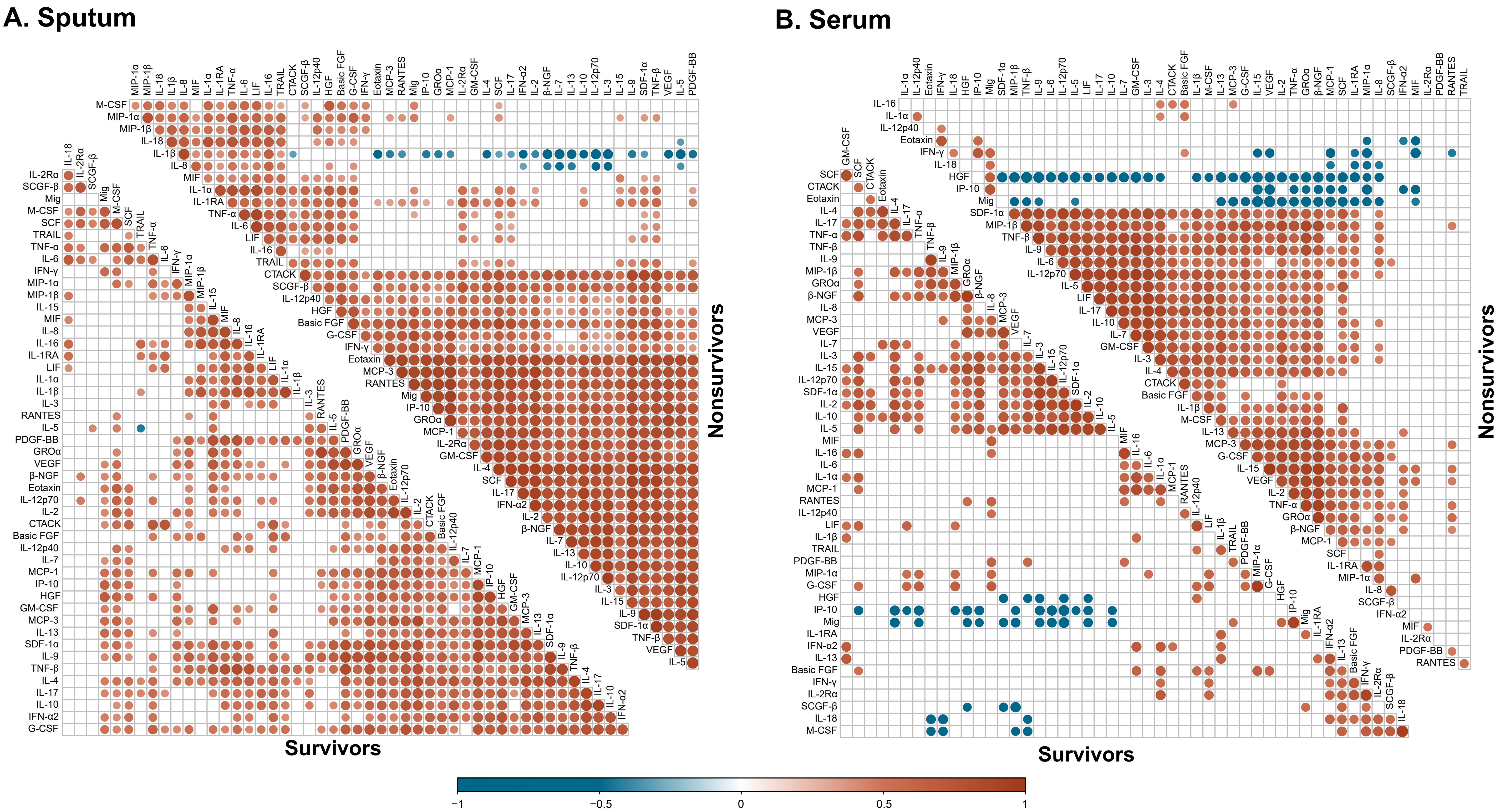


**Supplementary Figure 6. Correlation matrix across all time points of 48 cytokines in sputum/serum from five H5N6 patients with different outcomes**

**Legends of supplementary figures**

**Supplementary Figure 1. Key events timeline of five patients infected with influenza A(H5N6) virus in Guangdong Province, China, 2021-2022.**

Different filling colors represented various key events throughout disease courses. Position of positive sputum and blood cultures referred to the collection date of sputum and blood that had bacteria cultured.

*For patient 5, from D61 to D113, no special events except recurrent and refractory low $PaO_2/FiO_2$ ratio arose.

Abbreviation: $PaO_2$, partial pressure of oxygen in arterial blood; $FiO_2$, fraction of inspired oxygen.

**Supplementary Figure 2. Phylogenetic trees of HA and NA genes of influenza A (H5N6) viruses in five patients in Guangdong Province, 2021 to 2022.**

Phylogenetic trees of full-length HA (A) and NA (B) genes of H5N6 influenza A viruses derived from respiratory tract of five patients in Guangdong Province, China, 2021-2022.

Abbreviations: HA, hemagglutinin; NA, neuraminidase.

**Supplementary Figure 3. Correlations of $PaO_2/FiO_2$ ratio and SOFA scores with sputum viral load in five H5N6 patients.**

A, C, E, G and I, Correlation of $PaO_2/FiO_2$ ratio with viral load in sputum collected at the same date of $PaO_2/FiO_2$ evaluation. B, D, F, H and J, Correlation of SOFA scores with viral load in sputum collected at the same date of SOFA evaluation. *r* values and *P* values were determined by Spearman's correlation tests. *r* values reflect degrees of linear correlation between two variables and *P* values reflect the likelihood of correlation. *r* values range from -1 to 1: $r<0$, negative linear correlation; $r=0$, no linear correlation; $r>0$, positive linear correlation. With *P* values less than 0.05, there is likely a relation between changes of two variables.

**Supplementary Figure 4. Dynamic change of sputum cytokine concentrations in H5N6 patients over time.**

Dots represented individual data at different time points, with raw values plotted on the $\log_{10}$ scale. Different shapes of dots represented sputum cytokines of different patients.

**Supplementary Figure 5. Dynamic change of serum cytokine concentrations in H5N6 patients over time.**

Dots represented individual data at different time points, with raw values plotted on the $\log_{10}$ scale. Different shapes of dots represented serum cytokines of different patients.

**Supplementary Figure 6. Correlation matrix across all time points of 48 cytokines in sputum/serum from five H5N6 patients with different outcomes.**

Correlation matrix across all time points of 48 cytokines from patients' serum and sputum, comparing survivors with nonsurvivors. Correlations between sputum cytokines (A) and serum cytokines (B), respectively. Upper graphs indicated serum or sputum cytokines from nonsurvivors, bottom graphs indicated those from survivors. Blank grid meant no statistical significance. Color intensity and size of the circle are proportional to the correlation coefficients. Significance was determined by Spearman's correlation tests. A *P* value less than 0.05 was considered statistically significant.
